# Comparison of clinical course and outcomes of critically ill patients with SARS-CoV2 infection managed in traditional ICU and “Flex” ICU during the surge of the pandemic in the Bronx

**DOI:** 10.1101/2021.03.03.21252868

**Authors:** Victor Perez Gutierrez, Alex Carlos, Jessica Nino, Julieta Osella, Vihren Dimitrov, Moiz Kasubhai, Vidya Menon

## Abstract

**BACKGROUND:** As part of the response to increase critical care capacity during the unprecedented surge of COVID-19 infections, NYC Health + Hospital systems identified and resourced areas in the hospital that could deliver critical care as “Flex” ICUs to complement the traditional ICUs to manage the rapid influx of critically ill patients.

**OBJECTIVE:** Comparison of clinical features and outcomes of mechanically ventilated COVID-19 patients admitted to the traditional and “Flex” ICUs during the surge of the pandemic

**METHODS:** Retrospective comparative cohort study of patients with confirmed SARS-CoV-2 infection on mechanical ventilation admitted to traditional ICU and ‘Flex’ ICU. Univariate and multivariate analysis to detect factors associated with death from COVID-19 patients in mechanical ventilation were performed with the Cox proportional hazards regression model

**RESULTS:** Out of the 312 patients on mechanical ventilation, 111 were admitted to the traditional ICU and 201 to the ‘Flex’ ICU. The mortality rate was higher in the ‘Flex’ ICU compared with the traditional ICU, but the adjusted risk model was not significantly associated with increased mortality

**CONCLUSION:** **“**Flex” ICUs played a crucial role in the management of critically ill patients during the pandemic. Mortality risk of patients in the “Flex” ICUs were comparable to traditional ICUs in the adjusted analysis. While there is enough evidence for Intensivist managed ICUs to have better outcomes, our study demonstrates the feasibility of non-intensivist led Flex” ICUs during a crisis.

## INTRODUCTION

An already strained US critical care system was overwhelmed during the peak of the pandemic, as anticipated by experts based on experience from China and Italy(1). Initial reports from China suggested that approximately 5% of proven COVID-19 infections required intensive care(2); however, in the United States, as per the CDC report in March 2020, the hospitalization rate was 20.7-31.4% with 4.9-11.5% requiring intensive care(3,4). As New York City became the epicenter of the COVID-19 pandemic in March 2020, all city hospitals faced unprecedented stresses to ramp up capacity. With the surge of positive cases requiring hospitalization, the hospitals needed to increase capacity in the Emergency departments, Intensive care units, and regular units at an urgent and fast pace and scale. The NYC Health + Hospitals (NYC H+H) system is the largest provider of health services to the low-income and minority populations in the city. With these communities being disproportionately affected by the pandemic, the NYC H+H faced a unique challenge. The rapidly evolving clinical, operational needs across the eleven-hospital system were recognized, and efforts to increase capacity and resources were implemented. Critical care capacity was expanded system wide, with an increase from 300 bed ICU capacity at baseline to over 1000 beds. This was possible by increasing formal critical care beds, use of nontraditional hospital space creatively, increase adequate staffing, and supplement the supply of necessary equipment(5). Additional areas in the hospital that potentially had the logistics to deliver critical care were identified as “flex” ICU spaces. Optimum utilization of the available space was done by using oxygen “splitters,” retrofitting rooms with windows using High-Efficiency Particulate Air (HEPA) filters vented externally to create negative pressure rooms to increase capacity while following infection control requirements(5). The increase in number of patients with severe/critical COVID also placed a significant strain on staffing, especially ICU staffing across H+H. In addition to the redeployment of staff from suspended services like ambulatory care, a tiered staffing structure using intensivists and ICU nurses guided the non-ICU trained providers and increased ability to provide high-quality critical care. The system partnered with the US Department of Defense and engaged volunteers and private staffing agencies to help meet the demand of qualified personnel(6). While traditional ICUs managed by intensivists have been well known to deliver better outcomes, we examined the comparison of clinical features and outcomes of severe/critical COVID-19 patients requiring intensive care admitted to the traditional and “Flex” ICUs during the surge of the pandemic.

## METHODS

### Study design and inclusion criteria

This single center, retrospective, observational study was done at the NYC H+H in the Bronx. Patients with confirmed COVID-19 infection requiring ventilatory support and critical care from March 23, 2020, to May 1, 2020 (surge period for the hospital) were included in the study. As per the workflow during the surge, all mechanically ventilated patients were evaluated by the intensivist and triaged to ICU (traditional/flex). A confirmed case was defined as a positive nasopharyngeal swab by a real-time reverse transcription-polymerase test (Bio-Reference Laboratories, Inc., Elmwood Park, NJ, USA) for SARS CoV-2. Patients included in the study required mechanical ventilation and were ≥ 18 years old. Patients who were transferred intubated from a different Hospital or intubated for non-COVID related reasons as endoscopy were excluded from this study. Patients transferred to a different hospital or who died within 24 hours after intubation were excluded from the analysis (figure 1). All patients were followed until they expired or were discharged from our institution. The Traditional ICU was managed by intensivists (medical/surgical) with critical care nurses at a ratio of 1 for every 1-2 critically ill patients. The “Flex ICU” was managed by hospitalists/surgeons with critical care nurses teamed with general medical/surgical nurses at a ratio of 1 critical care nurse for 4 mechanically ventilated patients. The Institutional Review Board approved the study (IRB #20-007).

**Figure 1.**
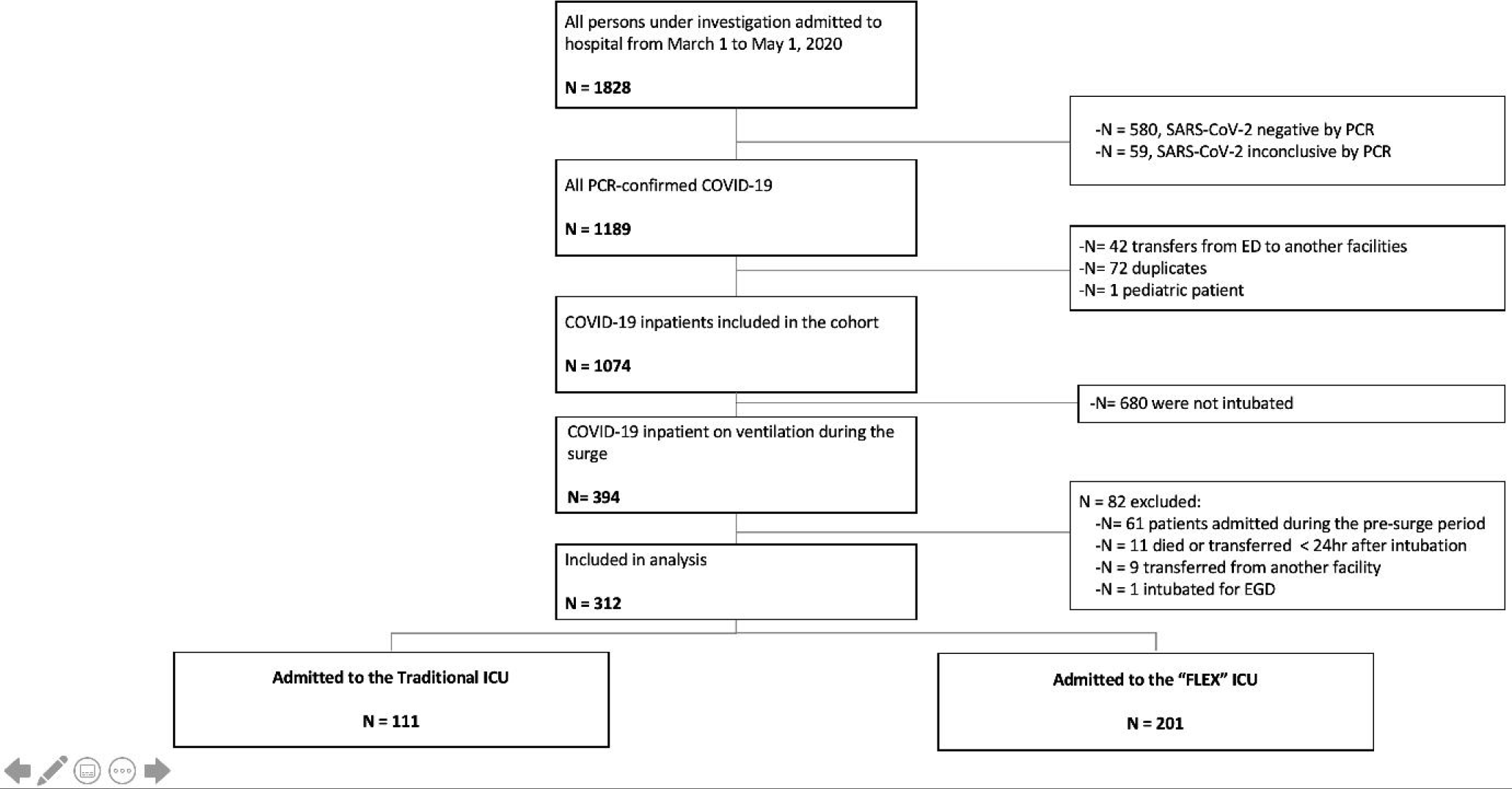
Flow diagram of process for patient inclusion as participant of the study

### Data collection and outcomes definition

Sociodemographic and broad characteristics including age, sex, ethnicity, and comorbidities categorized into hypertension, diabetes, chronic Lung Disease, chronic kidney disease, congestive heart disease, chronic liver disease and viral hepatitis, cancer, and HIV and presenting symptoms were collected from medical records. The Charlson Comorbidity Index (CCI) which predicts 10-year was used as a baseline characteristic and for adjustment as a potential confounding factor(7). Patients were stratified as Moderate, Severe, and Critical COVID-19 infection on admission-based WHO guidelines(8). Sepsis was defined based on qSOFA score (altered mental status, respiratory rate ≥22, and or systolic blood pressure ≤100)(9). The Berlin Definition of Acute Respiratory Distress Syndrome was applied to define ARDS on Admission(10). Acute Physiology and Chronic Health Evaluation (APACHE II) was performed within 24 hours after admission in ICU or FLEX unit(11). Compliance with mechanical ventilation standards like ARDS protocol as defined by NHLBI ARDS Network and Tidal volume delivery by ideal body weight were evaluated for 14 days after intubation(12). All clinical outcomes such as mortality, discharge, Length of stay, days of mechanical ventilation, extubations, and tracheostomy placement were collected. Ventilation-free days at 28 days were calculated for successfully weaned patients from mechanical ventilation within 28 days(13). Details of treatments (HCQ, Remdesivir, steroids, therapeutic anticoagulation) and complications affecting the course of hospitalization, including Acute kidney injury, requirement of renal replacement therapy, vasopressors, etc. were also obtained.

### Statistical analysis

Categorical variables were presented as frequencies and continuous variables as median (interquartile range [IQR]). Chi-square or the Fisher exact test was used to analyze categorical variables, and the Mann-Whitney test was applied to analyze non-normally distributed continuous variables. Kaplan-Maier survival curve and Long-rank test were performed in this study. Univariate and multivariate analysis to detect factors associated with death was performed by conducting the Cox proportional hazards regression model. Hazard ratio (HR) and 95% confidential interval (95% CI) were assessed. All reported *p* values were considered significant if <0.05. Data were analyzed using SPSS 26 (SPSS, Inc, Chicago, Ill).

## RESULTS

Of the 1074 patients hospitalized with confirmed COVID-19 infection between March 1^ST^-May 1^st^, 394 patients required invasive mechanical ventilation. Our study included 312 patients with positive SARS-CoV2 PCR who required mechanical ventilation (Figure 1), of which 111 (36 %) were admitted to the Traditional Intensive Care Unit and 201 (64%) to “FLEX” intensive units (figure 1).

Patients admitted in the ICU and FLEX were comparable concerning baseline characteristics (table 1).

**Table 1:** Baseline demographic and clinical presentation for Mechanically ventilated patients with COVID-19 infection.

|  |  |  |  |
| --- | --- | --- | --- |
|  | **Overall** | **Conventional ICU** | **“Flex” ICU** |
|  | **n=312** | **n=111** | **n=201** |
| Broad characteristics, No. (%) |  |  |  |
| Age, years (median, IQR) | 63 (54.0-73.0) | 62.0 (51.0-72) | 64.0 (54.5.0-73.0) |
| Age (years), No. (%) |  |  |  |
| <41 years | 26 (8.3%) | 16 (14.4%) | 10 (5.0%) |
| 41-60 years | 108 (34.7%) | 35 (31.5%) | 73 (36.3%) |
| 61-80 years | 148 (47.4%) | 52 (46.8%) | 96 (47.8%) |
| >80 years | 30 (9.6%) | 8 (7.2%) | 22 (10.9%) |
| Sex, female, No. (%) | 122 (39.1%) | 42 (37.8%) | 80 (39.8%) |
| Race, No. (%) |  |  |  |
| LatinX | 214 (68.6%) | 76 (68.5%) | 138 (68.7%) |
| Black | 84 (26.9%) | 31 (27.9%) | 53 (26.4%) |
| White | 8 (2.6%) | 1 (0.9%) | 7 (3.5%) |
| Asian | 6 (1.9%) | 3 (2.7%) | 3 (1.5%) |
| Body mass index, kg/m2 (median, IQR) | 31.0 (26.7-36.9) | 31.6 (27.7-38.0) | 301 (26.1-35.8) |
| Body mass index, No. (%) |  |  |  |
| < 18.5 kg/m2 | 4 (1.3%) | 0 (0.0%) | 4 (2.0%) |
| 18.5-24.9 kg/m2 | 41 (13.1%) | 11 (9.9%) | 30 (14.9%) |
| 25-29.9 kg/m2 | 95 (30.4%) | 32 (28.8%) | 63 (31.3%) |
| 30-39.9<br>kg/m2 | 125 (40.1%) | 47 (42.3%) | 78 (38.8%) |
| >40kg/m2 | 47 (15.1%) | 21 (18.9%) | 26 (12.9%) |
| Comorbidities, No. (%) |  |  |  |
| Diabetes | 142 (45.5%) | 52 (46.8%) | 90 (44.8%) |
| Hypertension | 150 (48.1%) | 54 (48.6%) | 98 (48.8%) |
| Obstructive lung disease (asthma or COPD) | 60 (19.2%) | 21 (18.9%) | 39 (19.4%) |
| Chronic kidney disease (Stage $\geq 3$) | 40 (12.8%) | 10 (9.0%) | 30 (14.9%) |
| Congestive heart failure | 20 (6.4%) | 7 (6.3%) | 13 (6.5%) |
| Cancer | 15 (4.8%) | 4 (3.6%) | 11 (5.5%) |
| Human immunodeficiency virus infection | 5 (1.6%) | 2 (1.8%) | 3 (1.5%) |
| Charlson Comorbidity Index (median, IQR) | 3.0 (1.0-5.0) | 3.0 (1.0-4.0) | 3.0 (2.0-5.0) |
| Clinical symptoms on presentation, No. (%) |  |  |  |
| Shortness of breath | 258 (82.7%) | 94 (84.7%) | 164 (81.6%) |
| Cough | 223 (71.5%) | 83 (74.6%) | 140 (69.7%) |
| Fever | 196 (62.8%) | 68 (61.3%) | 128 (63.7%) |
| Neurological symptoms (altered mental status) | 68 (21.8%) | 21 (18.9%) | 47 (23.4%) |
| GI symptoms (diarrhea or vomiting) | 52 (16.7%) | 17 (15.3%) | 35 (17.4%) |
| Severity of the disease, No. (%) |  |  |  |
| Severity of COVID-19 on admission |  |  |  |
| Moderate | 20 (6.4%) | 11 (9.9%) | 9 (4.5%) |
| Severe | 97 (31.1%) | 35 (31.5%) | 62 (30.8%) |
| Critical | 195 (62.5%) | 65 (58.6%) | 130 (64.7%) |
| Supplemental Oxygen required on admission (FiO2) | 1.0 (0.8-1.0) | 1.0 (0.8-1.0) | 1.0 (0.8-1.0) |
| APACHE-II score (median, IQR) at intubation | 14 (10-20) | 13 (10-20) | 14 (11-20) |
| ARDS on admission | 195 (62.5%) | 65 (58.6%) | 130 (64.7%) |
| Sepsis on admission by Quick SOFA | 187 (59.9%) | 69 (62.2%) | 118 (58.7%) |
| Radiological characteristics, No. (%) |  |  |  |
| Bilateral reticulonodular opacities | 204 (65.4%) | 75 (67.6%) | 129 (64.2%) |
| Ground-glass opacities | 106 (33.9%) | 32 (28.8%) | 74 (36.8%) |
| Focal consolidation | 38 (12.2%) | 12 (10.8%) | 26 (12.9%) |
| Atelectasis | 11 (3.5%) | 5 (4.5%) | 6 (3.0%) |
| Clinical course, No. (%) |  |  |  |
| PaO<sub>2</sub>/Fio<sub>2</sub> ratio before intubation | 116.4 (78.9-185.3) | 120 (77.8-187.9) | 115.0 (79.2-177.1) |
| >300 | 22 (7.1%) | 11 (9.9%) | 11 (5.9%) |
| 299-200 | 45 (14.4%) | 15 (13.5%) | 30 (14.9%) |
| 199-100 | 116 (37.2%) | 37 (33.3%) | 79 (39.3%) |
| <100 | 129 (41.3%) | 48 (43.2%) | 81 (40.3%) |
| BiPAP prior to mechanical ventilation | 38 (12.2%) | 14 (12.6%) | 24 (11.9%) |
| Vasopressors use during hospital course | 250 (80.1%) | 90 (81.1%) | 160 (79.6%) |
| Acute kidney injury during hospital course | 198 (63.5%) | 77 (69.4%) | 121 (60.2%) |
| Renal replacement therapy received | 42 (13.5%) | 20 (18.0%) | 22 (10.9%) |
| Bacterial superinfection | 33 (10.6%) | 16 (14.4%) | 17 (8.5%) |
| ARDS protocol followed 14 days | 42 (13.5%) | 16 (14.4%) | 26 (12.9%) |
| Tidal Volume by IBW followed 14 days | 63 (20.2%) | 26 (23.4%) | 37 (18.4%) |
| Barotrauma | 23 (7.4%) | 7 (6.3%) | 16 (8.0%) |
| Abbreviations: ARDS, acute respiratory distress syndrome; BiPAP, bilevel positive airway pressure; COPD, chronic obstructive pulmonary disease |  |  |  |

While the incidence of Acute respiratory distress syndrome on admission was 59% among patients admitted to traditional ICU, it was 65% among those in the Flex ICU cohort. The stratification of the ARDS between traditional and Flex ICU was Mild ARDS (13% vs. 14%), moderate ARDS (33% vs. 37%), and severe ARDS (43% vs. 41%) (Table 1). The Apache II score within 24 hours on intubation was comparable between the two groups (median, 13 vs. 14). The majority of patients in both cohorts required vasopressor drugs during hospital course (81% vs. 80%), developed acute kidney injury (69% vs. 60%), and had superimposed bacteremia (14% vs. 8%). Among patients with acute kidney injury, 18% from the traditional ICU cohort received Renal replacement therapy compared to 11% from the Flex ICU. In both units, compliance with ARDS protocol (14% vs. 13%) was similar, as was delivery of, tidal volume by Ideal body weight (23% vs. 18%). The use of therapeutic interventions like Hydroxychloroquine, azithromycin, Convalescent Plasma, therapeutic anticoagulation, and stress doses of corticosteroids was also similar in both cohorts. A similar percentage of patients in the traditional ICU and Flex ICU cohort underwent “Proning” as per our institutional protocol (Table 2).

**Table 2:** Treatment, intervention, and outcomes for Mechanically Ventilated patients with COVID-19 infection.

|  | Overall | Conventional ICU | “FLEX” intensive Unit | *p-value* |
| --- | --- | --- | --- | --- |
|  | n=312 | n=111 | n=201 |  |
| **Treatment and interventions, No. (%)** |  |  |  |  |
| **Hydroxychloroquine** | 271 (86.9%) | 97 (87.4%) | 174 (86.6%) |  |
| **Remdesevir** | 5 (1.6%) | 4 (3.6%) | 1 (0.5%) |  |
| **Convalescence Plasma** | 26 (8.3%) | 10 (9.0%) | 16 (8.0%) |  |
| **Azithromycin** | 281 (90.1%) | 100 (90.1%) | 181 (90.0%) |  |
| **Therapeutic Anticoagulation** | 121 (37.5%) | 46 (41.1%) | 75 (37.3%) |  |
| **Stress doses corticosteroids** | 88 (28.2%) | 36 (32.4%) | 52 (25.9%) |  |
| **Proning per protocol** | 87 (27.8%) | 34 (30.6%) | 53 (27.9%) |  |
| **Outcomes (followed 90 days), No. (%)** |  |  |  |  |
| **Deceased** | 245 (78.5%) | 76 (68.5%) | 169 (84.1%) | 0.04 |
| **Time to death (median, IQR) from admission** | 9.4 (6.2-15.1) | 9.0 (5.5-16.7) | 9.4 (6.7-14.9) | 0.43 |
| **Length of stay, days (median, IQR)** | 11.02 (7.0-19.0) | 11.7 (6.2-22.0) | 10.2 (7.0-17.7) | 0.42 |
| **Length of mechanical ventilation, days (median, IQR)** | 8.6 (5.1-16.0) | 9.0 (5.0-18.0) | 8.2 (5.2-14.0) | 0.42 |
| **Ventilator-free days (median, IQR) \*** | 18.5 (9.0-25.0) | 18.0 (10.0-25.0) | 19 (10.0-25.0) | 0.83 |
| **Total extubations** | 53 (17.0%) | 24 (21.6%) | 29 (14.4%) | 0.10 |
| **Terminal extubations** | 12 (3.8%) | 5 (4.5%) | 7 (3.5%) | 0.65 |
| **Successful extubations** | 41 (13.2%) | 19 (17.7%) | 22 (10.9%) | 0.12 |
| **Tracheostomy** | 40 (12.8%) | 22 (19.8%) | 18 (9.0%) | 0.03 |
| *Vent Free days was calculated by removing patients who died in less than 28 days or intubated more than 28 days. |  |  |  |  |

There was a significant difference (69% vs. 84%, p<0.05) in mortality between the traditional ICU and Flex ICU. However, no statistical difference was found in terms of time to death (median, 9.0 vs. 9.4), length of stay (11.7 vs. 10.2), length on mechanical ventilation (median, 9.0 vs. 8.2), and ventilation-free days (median, 18.0 vs. 19.0). Total extubations were comparable in both units (21.6% vs 14.4%) and tracheostomy were more frequent in ICU (19.8% vs 9.0%, p<0.05).

Kaplan-Meier survival analysis to compare the effectiveness in preventing death between the traditional Intensive care unit and Flex unit is shown in Fig 2. Subjects managed in traditional ICU had a median time to survive of 13.54 (95% CI, 8.74 to 18.33) days, which was longer than the Flex ICUs group 10.60 (95% CI, 9.16-12.09) days. Significant differences were found in the survival distribution for Long Rank tests (p<0.05).

**FIGURE 2:**
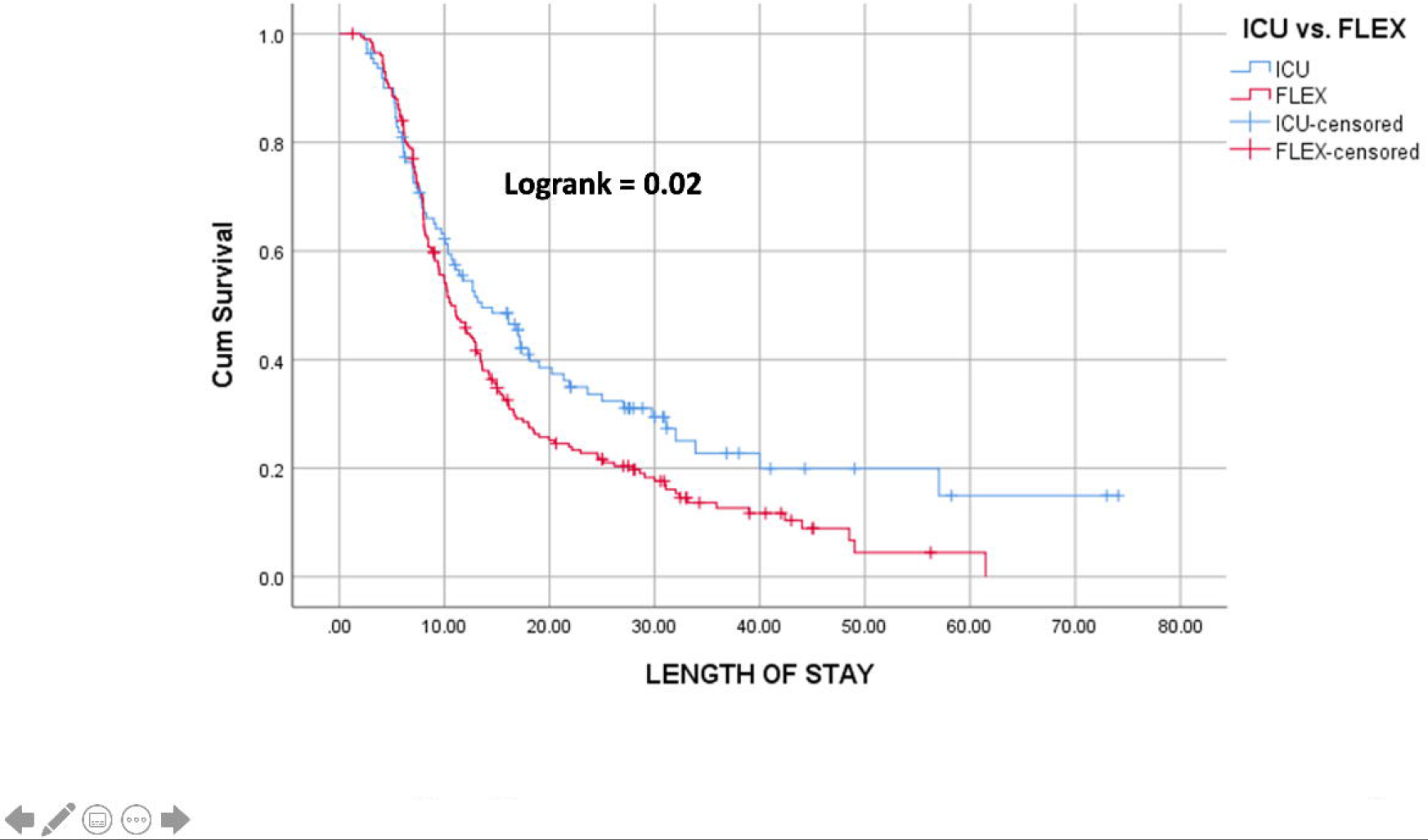
KAPLAN MEIER SURVIVAL CURVE FOR MECHANICALLY VENTILATED PATIENT WITH COVID-19 INFECTION BY UNIT. Kaplan-Meier survival analysis to compare the effectiveness in preventing death between the traditional Intensive care unit and Flex unit is shown in Fig 2. Significant differences were found in the survival distribution for Logrank tests (p=0.02).

On Univariate cox proportional hazard regression analysis, the mortality risk in the ‘Flex’ ICU was significantly higher than the Traditional ICU. However, following covariate adjustment, the risk decreases and becomes comparable (adjusted, HR, 1.28, 95% CI, 0.97-1.70, p=0.078). The covariates, age 65 years or more (adjusted HR, 1.65, 95% CI, 1.25-2.17, p<0.001), Latinx (Adjusted HR, 1.33, 95% CI, 1.01-1.761, p<0.05), Critical status of patients with COVID-19 infection on admission (Adjusted, 1.90, 95 % CI, 1.01-2.17, p<0.05) and sepsis syndrome by qSOFA score (Adjusted, HR, 1.57, 95% CI, 1.20-2.05, p<0.05) were independent risk factors associated with death in mechanical ventilated patient with COVID-19 infection (table 3).

**Table 3:** Univariable and Multivariable Cox Proportional Hazzard Ratio for Mortality in Mechanically Ventilated patients with COVID-19 infection.

|  | Univariable |  |  | Multivariable |  |  |
| --- | --- | --- | --- | --- | --- | --- |
|  | HR | 95 % CI | p-value | HR | 95 % CI | p-value |
| **UNIT** |  |  |  |  |  |  |
| Conventional ICU | Reference |  |  | Reference |  |  |
| “FLEX” UNIT | 1.37 | 1.05-1.81 | <0.05 | 1.29 | 0.97-1.71 | 0.078 |
| **Age (>65yo vs <65yo)** | 1.62 | 1.26-2.09 | <0.001 | 1.65 | 0.51-.088 | 0.001 |
| **Female** | 0.89 | 0.69-1.15 | 0.40 | 0.77 | 0.58-1.01 | 0.070 |
| **Race** |  |  |  |  |  |  |
| **Non-Latinx** | Reference |  |  | Reference |  |  |
| **Latinx** | 1.28 | 0.97-1.67 | 0.07 | 1.34 | 1.01-1.76 | 0.035 |
| **BMI\*($>30 \text{ kg/m}^2$ vs $<30 \text{ kg/m}^2$)** | 1.12 | 0.87-1.44 | 0.37 | 1.05 | 0.81-1.37 | 0.700 |
| **Hypertension** | 0.97 | 0.75-1.23 | 1.23 | 0.81 | 0.61-1.07 | 0.130 |
| **Diabetes** | 1.16 | 0.90-1.50 | 0.22 | 1.18 | 0.89-1.56 | 0.250 |
| **Chronic kidney disease** | 1.24 | 0.87-1.79 | 0.22 | 1.09 | 0.75-1.59 | 0.700 |
| **COVID severity** |  |  |  |  |  |  |
| **Moderate** | Reference |  |  | Reference |  |  |
| **Severe** | 1.79 | 0.94-3.38 | 0.70 | 1.53 | 0.80-2.94 | 0.200 |
| **Critical** | 2.05 | 1.11-3.78 | $<0.05$ | 1.91 | 1.02-2.06 | 0.030 |
| **Sepsis syndrome\*** | 1.53 | 1.18-2.00 | $<0.001$ | 1.58 | 1.21-2.06 | 0.030 |
| **APACHE II score** | 0.94 | 0.73-1.22 | 0.70 | 1.01 | 0.99-1.03 | 0.290 |
| *BMI, body mass index; *Sepsis syndrome defined by Quick SOFA score. |  |  |  |  |  |  |

## DISCUSSION

It is well established that critically ill patients have decreased mortality and length of stay when managed by trained intensivists(14– 16). Due to the unprecedented demand for critical care services at the peak of the pandemic, hospitalists and surgeons stepped up to provide intensive care in the designated “Flex” ICUs. Our study presents the comparison of patient outcomes between a traditional ICU and Flex ICU. While both groups were comparable, it was not surprising that the survival outcomes were better in the traditional ICUs with shorter lengths of stay. Notably, the Flex ICUs managed a higher proportion of patients older than 80 years and those admitted with critical COVID-19 infection on admission. Though the Charlson Comorbidity indices between the two groups were comparable, the Flex ICUS managed more CKD patients than the traditional ICUs. While unadjusted outcomes in the traditional ICUs were significantly better than the Flex units, the risk was reduced to become almost comparable on the adjustment model. Notably, the patients anticipated to have better outcomes (younger age, fewer comorbidities) as per intensivists assessments following intubation and initiation of mechanical ventilation, were appropriately triaged to the traditional ICUs to deliver best outcomes.

NYC’s public health system, NYC Health +Hospital, expanded capacity across its eleven acute care facilities and added three new field hospitals in response to the surge of COVID-19 infection in the city. As per the direction of the senior administration of the system, all of the system hospitals with their 4000 total beds joined and prepared to become a single large intensive care unit(5,17). As part of this response our hospital, increased capacity from a 34 bed ICU capacity to 195 beds by innovatively repurposing nontraditional hospital space and equipment with staff support from the US Department of Defense, private staffing agencies, and volunteer medical staff from all over the country. An important consideration also was allocation of staffing to handle the increased volume and severity of illness of patients. After focusing on evidence-based education and retraining in record time, the existing staff and new hires were deployed to critical care. That being said, the nurse staffing in the Flex ICUs was different from the traditional ICUs, with a ratio of 4 patients to one critical care nurse in comparison to two patients to one nurse in the traditional ICUs. The flex ICUs also had med-surg nurses to manage the patients, not requiring mechanical ventilation, who assisted the critical care nurses in managing the ventilated patients with medication management and other patient care responsibilities. The Flex ICUs had limited direct visibility and telemetry monitoring of critically ill patients. There were additional constraints due to stretching of services of respiratory therapists and other support staff in the flex-ICUs compared to traditional ICUs with established systems, staffing, and protocols. Another major issue was the availability of mechanical ventilators. As the demand increased, the network supply of ventilators in the system was supplemented with ventilators from the federal stockpile. While the traditional ICUs received the standard ventilators, the flex-ICUs managed predominantly with transport ventilators, which are suboptimal to manage ARDS. We believe the in spite of our best efforts; the above-mentioned factors could have had an impact on outcomes.

Overall, life expectancy is lower in the South Bronx as compared to other NYC boroughs due to the high incidence of chronic health conditions as diabetes, hypertension, and chronic kidney disease(18,19). Few of the barriers faced by the community contributing to the delay in seeking timely help include, majority (58%) of the population being LatinX with Spanish as their primary language, a significant percentage of people living with an undocumented/undomiciled status without health insurance and a tendency to delay seeking medical attention till severely compromised(20). Notably, 37% of patients with COVID-19 admitted to our facility required mechanical ventilation and critical care compared to 13-28% reported in other studies from NYC(21,22).

The differences in mortality could thus be explained by a multitude of system and patient-related factors.

The rapid influx of patients with different stages of severity of the disease, evolving knowledge, and evidence-based clinical therapeutics, the challenge of reducing transmission of this infection over time, plus an unequal distribution of the human resource and supply chain played an essential role in outcomes.

Going forward with lessons learned from this experience, it is valuable to build capacity with respect to critical care staffing, cross-training physicians, nurses, and allied health specialists for intensive care, and map out a workflow to increase beds including ICU beds and source supplies to respond to the next wave of infections.

## CONCLUSION

The Flex ICUs created to handle the surge of critically ill COVID −19 patients were managed predominantly by non-intensivists (hospitalists/surgeons). While the survival of patients in the traditional ICUs was better than the Flex ICUs on univariate analysis, following adjustment of covariates, the differences reduced and became almost comparable. In conclusion, while there is enough evidence for Intensivist managed ICUs to have better outcomes, it is possible to manage patients safely with good outcomes in the Flex ICU settings during a crisis with non-intensivists with training and adequate nursing/resource support.

## Data Availability

Data available as request

## REFERENCE

1. Blumenthal D, Fowler EJ, Abrams M, Collins SR. Covid-19 — Implications for the Health Care System. N Engl J Med. 2020 Oct 8;383(15):1483–8.

2. Zhou F, Yu T, Du R, Fan G, Liu Y, Liu Z, et al. Clinical course and risk factors for mortality of adult inpatients with COVID-19 in Wuhan, China: a retrospective cohort study. Lancet Lond Engl. 2020 28;395(10229):1054–62.

3. CDC. Coronavirus Disease 2019 (COVID-19) in the U.S. [Internet]. Centers for Disease Control and Prevention. 2020 [cited 2020 Oct 16]. Available from: https://covid.cdc.gov/covid-data-tracker

4. COVID-19: Data Main - NYC Health [Internet]. [cited 2020 Oct 16]. Available from: https://www1.nyc.gov/site/doh/covid/covid-19-data.page

5. A U Dm S, M S, S N, L B, Rj S, et al. Critical Care And Emergency Department Response At The Epicenter Of The COVID-19 Pandemic [Internet]. Vol. 39, Health affairs (Project Hope). Health Aff (Millwood); 2020 [cited 2020 Oct 11]. Available from: https://pubmed.ncbi.nlm.nih.gov/32525713/

6. Keeley C, Jimenez J, Jackson H, Boudourakis L, Salway RJ, Cineas N, et al. Staffing Up For The Surge: Expanding The New York City Public Hospital Workforce During The COVID-19 Pandemic. Health Aff Proj Hope. 2020 Aug;39(8):1426–30.

7. Charlson ME, Pompei P, Ales KL, MacKenzie CR. A new method of classifying prognostic comorbidity in longitudinal studies: development and validation. J Chronic Dis. 1987;40(5):373–83.

8. Clinical management of COVID-19 [Internet]. [cited 2020 Oct 16]. Available from: https://www.who.int/publications-detail-redirect/clinical-management-of-covid-19

9. Lambden S, Laterre PF, Levy MM, Francois B. The SOFA score— development, utility and challenges of accurate assessment in clinical trials. Crit Care [Internet]. 2019 Nov 27 [cited 2020 Oct 16];23. Available from: https://www.ncbi.nlm.nih.gov/pmc/articles/PMC6880479/

10. Force* Tadt. Acute Respiratory Distress Syndrome: The Berlin Definition. JAMA. 2012 Jun 20;307(23):2526–33.

11. Knaus WA, Draper EA, Wagner DP, Zimmerman JE. APACHE II: a severity of disease classification system. Crit Care Med. 1985 Oct;13(10):818–29.

12. NHLBI ARDS Network | About [Internet]. [cited 2020 Oct 16]. Available from: http://www.ardsnet.org/

13. Schoenfeld DA, Bernard GR, ARDS Network. Statistical evaluation of ventilator-free days as an efficacy measure in clinical trials of treatments for acute respiratory distress syndrome. Crit Care Med. 2002 Aug;30(8):1772–7.

14. Brown JJ, Sullivan G. Effect on ICU mortality of a full-time critical care specialist. Chest. 1989 Jul;96(1):127–9.

15. Hanson CW, Deutschman CS, Anderson HL, Reilly PM, Behringer EC, Schwab CW, et al. Effects of an organized critical care service on outcomes and resource utilization: a cohort study. Crit Care Med. 1999 Feb;27(2):270–4.

16. Fuchs RJ, Berenholtz SM, Dorman T. Do intensivists in ICU improve outcome? Best Pract Res Clin Anaesthesiol. 2005 Mar;19(1):125–35.

17. NYC Health + Hospitals to Triple ICU Capacity, Expand Personnel [Internet]. [cited 2020 Oct 16]. Available from: https://www.nychealthandhospitals.org/pressrelease/nyc-health-hospitals-to-triple-icu-capacity-expand-personnel/

18. Kaplan SA, Calman NS, Golub M, Davis JH, Ruddock C, Billings J. Racial and ethnic disparities in health: a view from the South Bronx. J Health Care Poor Underserved. 2006 Feb;17(1):116–27.

19. Newman D, Levine E, Kishore SP. Prevalence of multiple chronic conditions in New York State, 2011–2016. PLOS ONE. 2019 Feb 7;14(2):e0211965.

20. The Demographic Statistical Atlas of the United States - Statistical Atlas [Internet]. [cited 2020 Oct 16]. Available from: https://statisticalatlas.com/neighborhood/New-York/New-York/South-Bronx/Race-and-Ethnicity

21. Yehia BR, Winegar A, Fogel R, Fakih M, Ottenbacher A, Jesser C, et al. Association of Race With Mortality Among Patients Hospitalized With Coronavirus Disease 2019 (COVID-19) at 92 US Hospitals. JAMA Netw Open. 2020 Aug 3;3(8):e2018039.

22. Singer AJ, Morley EJ, Meyers K, Fernandes R, Rowe AL, Viccellio P, et al. Cohort of Four Thousand Four Hundred Four Persons Under Investigation for COVID-19 in a New York Hospital and Predictors of ICU Care and Ventilation. Ann Emerg Med. 2020;76(4):394–404.

